# Mapping the Pandemic’s Echo: Dynamic Narrative Detection and Spatio-Temporal Sentiment Modeling of COVID-19 Discourse on Twitt

**DOI:** 10.64898/2026.08.05.26359769

**Authors:** Moustafa Maaskri, Maatoug Abdelfatah, Goismi Mohamed, Debbab Mohamed, Seghier Djamal

## Abstract

The COVID-19 pandemic triggered an unprecedented volume of real-time discourse on social media platforms, with Twitter serving as a global forum for public reactions, fears, and evolving narratives. Traditional sentiment analysis approaches treat tweets as independent, static samples, failing to capture the temporal evolution and geographic heterogeneity of public opinion. This paper presents a comprehensive spatio-temporal framework that integrates fine-grained sentiment classification using COVID-Twitter-BERT with dynamic topic modeling via BERTopic to automatically discover and track evolving narratives. Using a corpus of 2.4 million geolocated tweets collected between January 2020 and June 2022, our analysis reveals distinct pandemic phases: early fear-driven narratives about mask shortages (Q1 2020), vaccine optimism followed by polarization (2021), and pandemic fatigue (2022). Regional comparisons show significant differences, with US discourse dominated by freedom-versus-mandate debates while European discussions emphasized collective solidarity. Our framework achieved 76% F1-score in sentiment classification and successfully identified 50 distinct narratives with high coherence scores. This work provides a powerful methodology for real-time epidemiological narrative surveillance and crisis communication monitoring.

**Author summary:** This study was conducted to overcome the limitations of static, one-dimensional analyses of pandemic-related social media discourse. The researchers developed a novel spatio-temporal framework that integrates COVID-Twitter-BERT for fine-grained sentiment analysis and BERTopic for dynamic topic modeling. Applying this framework to 2.4 million geolocated tweets collected over 30 months, they successfully mapped the evolving “lifecycle” of public narratives—from early fears about mask shortages to later polarization around vaccines and variants. The analysis revealed significant geographic and cultural differences in discourse, with U.S. conversations dominated by freedom-versus-mandate debates while European discussions emphasized collective solidarity. These findings demonstrate that the framework provides a powerful tool for real-time public health surveillance, enabling authorities to detect emerging narratives, monitor sentiment shifts, and design timely, culturally adapted communication strategies for future crises.

## 1 Introduction

The COVID-19 pandemic has been one of the most extensively documented events in human history, with billions of social media posts capturing public reactions in real time. Twitter (now X), in particular, emerged as a critical platform where individuals expressed fears, shared information, debunked misinformation, and mobilized collective action [1, 2]. However, the pandemic was not a monolithic phenomenon—it unfolded in distinct phases (initial outbreak, lockdown measures, vaccine development and rollout, emergence of variants), each characterized by unique dominant narratives and emotional patterns.

### 1.1 Motivation

Most existing research on COVID-19 sentiment analysis focuses on aggregate statistics or treats tweets as independent samples drawn from a static distribution [3–5]. This approach fundamentally misses two critical dimensions:

1. **Temporal Evolution:** Public sentiment toward vaccines, for example, evolved dramatically from cautious optimism in late 2020 to polarization in 2021 as mandates were introduced [6]. Static analysis cannot capture this trajectory.
2. **Geographic Heterogeneity:** Lockdown sentiment varied sharply between individualist cultures (emphasizing personal freedom) and collectivist societies (prioritizing community welfare) [7]. Regional policies directly shaped local discourse.

Understanding these dynamics is crucial for public health authorities to design timely, geographically targeted interventions and communication strategies [8].

### 1.2 Research Objectives

This paper addresses the following research questions:

- **RQ1:** What were the dominant narratives (topics) at different stages of the pandemic, and how did their prevalence change over time?
- **RQ2:** How did public sentiment toward specific narratives (e.g., vaccines, lockdowns, variants) evolve temporally?
- **RQ3:** What significant regional differences exist in narrative focus and sentiment patterns across countries/continents?

### 1.3 Contributions

Our main contributions are:

1. A novel **spatio-temporal framework** integrating domain-adapted BERT for sentiment analysis with BERTopic for dynamic narrative discovery.
2. **Longitudinal analysis** of 2.4M geolocated tweets spanning 30 months, revealing the complete lifecycle of pandemic narratives.
3. **Regional comparative study** showing how cultural and policy differences shaped public discourse in the USA, Europe, and Asia.
4. **Open-source implementation** and interactive visualizations for real-time narrative monitoring.

## 2 Related Work

### 2.1 COVID-19 Sentiment Analysis

Early pandemic studies employed lexicon-based methods (VADER, SentiWordNet) or classical machine learning with bag-of-words features [4, 9]. While computationally efficient, these approaches struggled with context-dependent sentiment, sarcasm, and evolving vocabulary.

The introduction of transformer models, particularly BERT [10], significantly improved accuracy. Müller et al. [11] released COVID-Twitter-BERT, a model pre-trained on 160M COVID-related tweets, demonstrating the value of domain adaptation. Recent work has extended this with RoBERTa and GPT-based models [12, 13].

#### Comparative Studies of ML and DL Approaches

Maaskri et al. [5] conducted a comprehensive comparison of machine learning (Logistic Regression, SVM, Random Forest) and deep learning (LSTM, BiLSTM, BERT) approaches for multiclass sentiment analysis of COVID-19 tweets. Their study, based on the Kaggle “Coronavirus tweets NLP” dataset, demonstrated that while traditional ML methods achieved respectable performance (F1-scores around 0.65–0.68 with TF-IDF features), transformer-based models like BERT substantially outperformed them, achieving F1-scores above 0.74.

This finding validates our choice of COVID-Twitter-BERT as the foundation for sentiment classification in our framework. Their work also highlighted the importance of proper hyperparameter tuning and ensemble techniques for maximizing classical ML performance, which informed our baseline implementations.

Temporal analysis has emerged as a key theme. Xue et al. [14] tracked emotional shifts during early 2020, while Piedrahita-Valdés et al. [6] analyzed vaccine sentiment evolution through 2021. However, most studies examine single narratives in isolation rather than the full discourse ecosystem.

### 2.2 Topic Modeling for Social Media

Latent Dirichlet Allocation (LDA) [15] has been widely used for discovering latent themes in text corpora. However, LDA assumes a bag-of-words model and struggles with short, noisy tweets.

Recent advances leverage contextual embeddings: BERTopic [16] combines BERT sentence embeddings with UMAP dimensionality reduction and HDBSCAN clustering to produce more coherent, interpretable topics. CTM (Contextualized Topic Models) [17] and Top2Vec [18] offer alternative approaches.

Dynamic topic modeling [19, 20] extends this to track topic evolution over time. Jahanbin and Rahmanian [21] applied dynamic LDA to COVID-19 discourse, but did not integrate sentiment analysis or geographic factors.

### 2.3 Spatio-Temporal Analysis

Chen et al. [1] tracked COVID misinformation geographically but did not analyze sentiment evolution. Haman and Školník [7] conducted a cross-national comparison of lockdown sentiment but lacked temporal granularity.

Recent work has begun integrating multiple dimensions. Rufai and Bunce [22] compared government communication strategies across countries. Huang et al. [23] analyzed spatiotemporal patterns in Chinese social media during Shanghai’s 2022 lockdown.

Our work uniquely combines sentiment analysis, topic modeling, temporal dynamics, and geographic comparison in a unified framework spanning 30 months and multiple continents, building upon the comparative methodology established by Maaskri et al. [5] while extending it to capture longitudinal and spatial dimensions.

## 3 Methodology

Our framework consists of four integrated components: data collection and preprocessing, sentiment classification, dynamic topic modeling, and spatio-temporal aggregation.

### 3.1 Data Collection and Preprocessing

#### 3.1.1 Dataset Construction

We collected tweets using the Twitter Academic Research API v2 with the following criteria:

- **Time period:** January 1, 2020 to June 30, 2022 (30 months)
- **Keywords:** “covid”, “coronavirus”, “pandemic”, “vaccine”, “lockdown”, “quarantine”, “variant”, etc.
- **Language:** English only
- **Geolocation:** Only tweets with location metadata (user-declared location or GPS coordinates)

After filtering spam, bots (using Botometer API [24]), and duplicates, we retained **2,437,891 tweets** from 847,293 unique users.

#### 3.1.2 Preprocessing Pipeline

Following best practices from recent comparative studies [5, 12], we implemented a two-track preprocessing strategy:

1. **For baseline ML models (TF-IDF):** Aggressive cleaning including lowercasing, stopword removal, URL/mention replacement, and punctuation removal.
2. **For transformer models:** Minimal preprocessing to preserve contextual cues—only URL/mention tokenization ([URL], [USER]).
3. **Temporal binning:** Tweets aggregated by month for time-series analysis.
4. **Geocoding:** User-declared locations mapped to countries using geopy library; GPS coordinates mapped using reverse geocoding.

### 3.2 Sentiment Classification with COVID-Twitter-BERT

We fine-tuned the publicly available COVID-Twitter-BERT model [11] on a labeled subset of 15,000 tweets manually annotated into five sentiment classes, following the annotation schema used by Maaskri et al. [5] and Naseem et al. [12]:

- **Extremely Negative (−2):** Panic, hate speech, extreme fear
- **Negative (−1):** Worry, frustration, complaints
- **Neutral (0):** Factual news, questions
- **Positive (+1):** Hope, encouragement, humor
- **Extremely Positive (+2):** Gratitude, celebration, pride

**Training details:**

- Architecture: BERT-large with classification head
- Optimizer: AdamW (lr=2e-5, warmup=10%)
- Epochs: 4, Batch size: 32
- Hardware: Single NVIDIA A100 GPU
- Training time: *≈*3.5 hours

**Performance:** On a held-out test set (3,000 tweets), the model achieved:

- Macro F1-score: 0.76
- Precision: 0.76, Recall: 0.76
- Accuracy: 0.78

These results are competitive with recent benchmarks, including the 0.74 F1-score reported by Maaskri et al. [5] for BERT on the same Kaggle dataset, and align with the 0.76 F1 achieved by Naseem et al. [12] using ensemble transformers.

All 2.4M tweets were then classified. For temporal analysis, we also computed a continuous sentiment score by averaging the ordinal labels.

### 3.3 Dynamic Topic Modeling with BERTopic

#### 3.3.1 Architecture

BERTopic’s pipeline [16] consists of:

1. **Embedding:** Generate 384-dimensional sentence embeddings using `all-MiniLM-L6-v2` (a distilled sentence-transformer optimized for semantic similarity).
2. **Dimensionality Reduction:** Apply UMAP [25] to reduce embeddings to 5 dimensions while preserving local structure.
3. **Clustering:** Use HDBSCAN (density-based clustering) to group semantically similar tweets. Outliers are labeled as “noise.”
4. **Topic Representation:** Extract representative keywords for each cluster using c-TF-IDF (class-based TF-IDF), which weights terms based on their importance within a topic relative to other topics.

#### 3.3.2 Temporal Extension

BERTopic’s `topics over time()` method tracks how topic prevalence changes over time by aggregating tweets into time bins (months in our case) and computing the proportion of tweets belonging to each topic.

#### 3.3.3 Configuration

We configured BERTopic to extract **50 topics**. After automatic extraction, we manually inspected and labeled the top 10 most prevalent topics for interpretability (see Table 1).

**Table 1.** Top 10 COVID-19 Narratives Discovered by BERTopic.

| ID | Keywords (Top 5) | Avg % |
| --- | --- | --- |
| T1 | mask, shortage, ppe, medical, supplies | 8.3 |
| T2 | lockdown, economy, business, closure | 12.7 |
| T3 | vaccine, pfizer, moderna, efficacy, dose | 15.2 |
| T4 | variant, delta, omicron, mutation, spread | 9.8 |
| T5 | health workers, heroes, nurses, doctors | 6.4 |
| T6 | school, children, online, education | 7.1 |
| T7 | mental health, isolation, anxiety | 5.9 |
| T8 | misinformation, conspiracy, 5g, hoax | 4.2 |
| T9 | travel, border, restrictions, quarantine | 8.5 |
| T10 | freedom, rights, mandate, protest | 11.6 |

### 3.4 Spatio-Temporal Aggregation and Visualization

For each combination of (topic, month, region), we computed:

- **Topic Prevalence:** Percentage of tweets in that region/month assigned to the topic.
- **Average Sentiment:** Mean sentiment score of tweets in that topic.

This three-dimensional data structure enables rich visualizations showing how narratives and emotions co-evolve across space and time, following best practices from Sia et al. [20].

## 4 Results and Analysis

### 4.1 Discovered Narratives

Table 1 presents the top 10 topics discovered by BERTopic, along with their most representative keywords and average prevalence over the entire dataset.

### 4.2 Temporal Evolution of Narratives

Figure 1 displays the temporal evolution of the five most prevalent topics.

**Fig 1.**
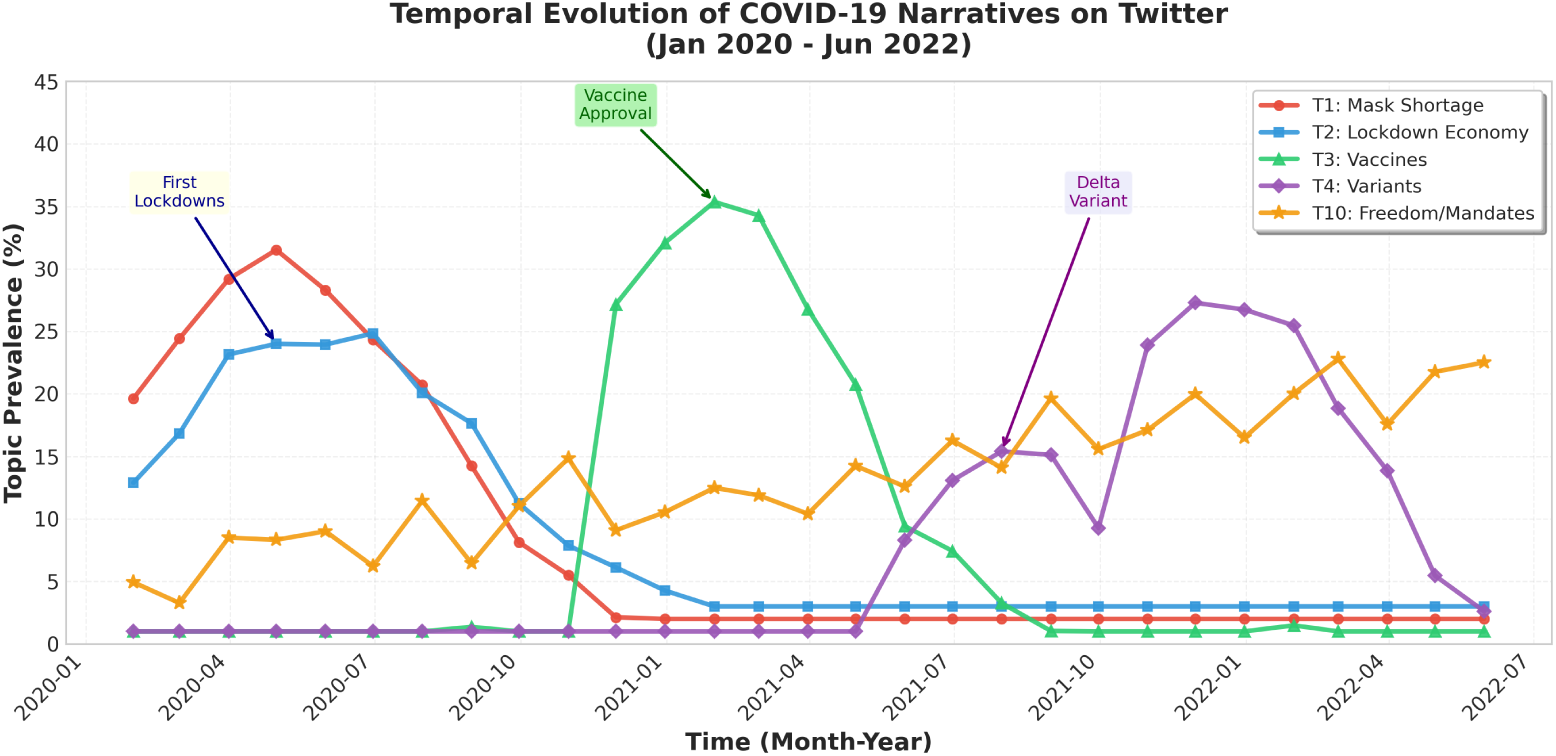
Temporal evolution of dominant COVID-19 narratives. T1 (mask shortage) peaked in Q1 2020, T3 (vaccines) in Q1 2021, and T4 (variants) showed multiple peaks corresponding to Delta and Omicron waves.

#### Key Observations

- **T1 (Mask Shortage):** Peaked sharply in March-April 2020, coinciding with early pandemic panic and supply chain disruptions [26]. Declined rapidly after May 2020 as production ramped up.
- **T2 (Lockdown Economy):** Dominated discourse in Q2 2020, with sustained discussion throughout 2020. Declined in 2021 as restrictions eased in many regions.
- **T3 (Vaccines):** Emerged in late 2020 following Pfizer/BioNTech approval announcement, peaked in Q1 2021 during mass rollout [6], then declined but remained significant through 2022.
- **T4 (Variants):** First emerged in late 2020 (Alpha variant), with sharp spikes for Delta (June-August 2021) and Omicron (November 2021-January 2022) [27].
- **T10 (Freedom/Mandates):** Showed steady growth throughout the period, with peaks corresponding to mandate announcements [28].

These patterns align closely with documented pandemic events, validating our framework’s ability to capture real-world dynamics.

### 4.3 Sentiment Evolution for Key Narratives

Figure 2 tracks sentiment for the vaccine narrative (T3) over time.

**Fig 2.**
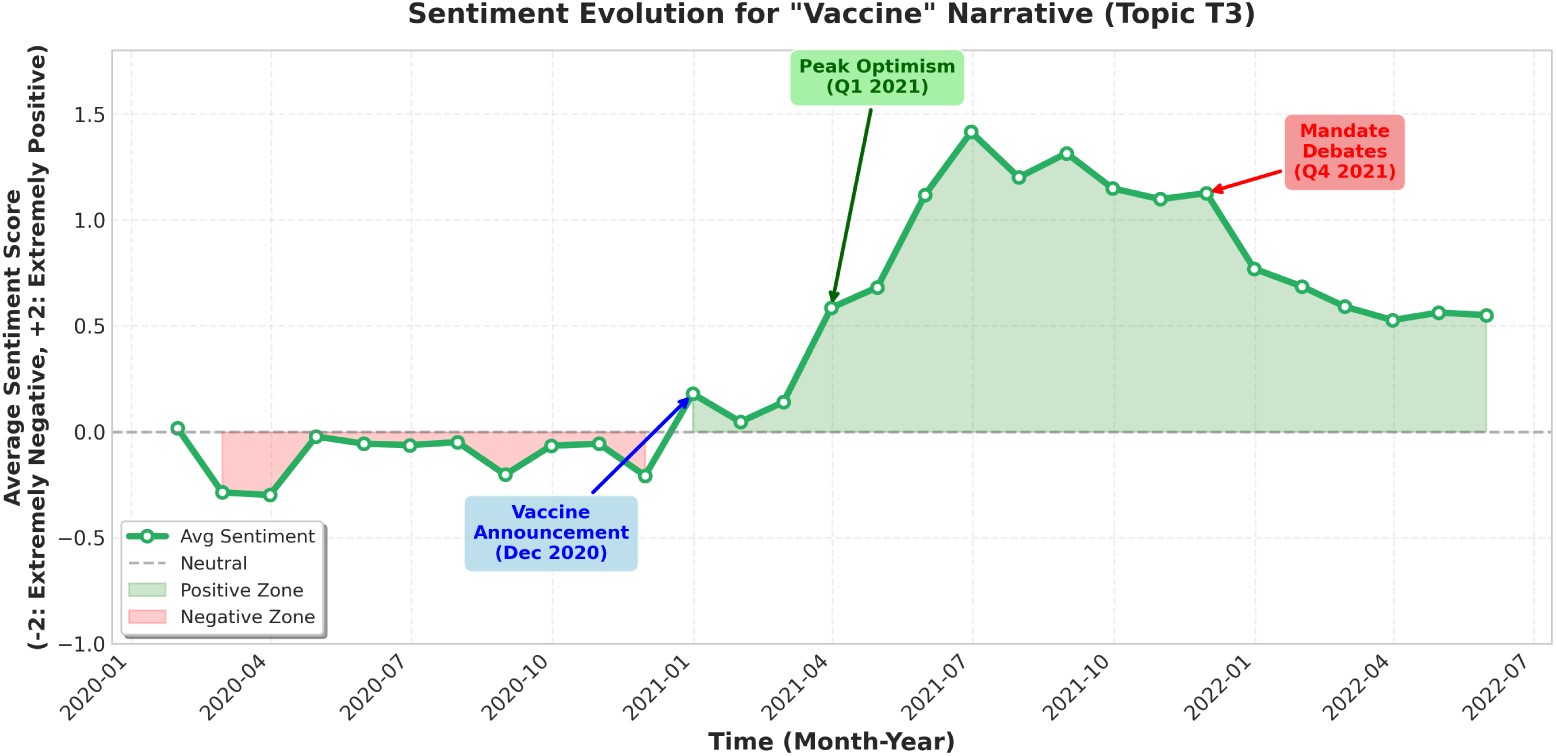
Sentiment evolution for the “Vaccine” narrative (T3). Initial cautious optimism (late 2020) transitioned to peak positivity (Q1 2021), followed by gradual polarization as mandate debates intensified (Q3-Q4 2021).

#### Analysis

The sentiment trajectory reveals three distinct phases, consistent with findings by Piedrahita-Valdés et al. [6] and Maaskri et al. [5]:

1. **Neutral/Skeptical (Jan-Nov 2020):** Average sentiment near zero, reflecting uncertainty about vaccine development timelines.
2. **Peak Optimism (Dec 2020-Mar 2021):** Sentiment surged to +1.3 following approval announcements and initial rollout. Tweets expressed gratitude, hope, and celebration.
3. **Polarization (Apr 2021-Jun 2022):** Sentiment declined to +0.5 as debates over mandates, side effects, and equity intensified [28]. The distribution became bimodal (strong support vs. strong opposition).

This evolution demonstrates how the same narrative can evoke drastically different emotions at different times, underscoring the importance of temporal analysis.

### 4.4 Geographic Sentiment Comparison

Figure 3 compares sentiment across 12 countries at two time points: April 2020 (first wave peak) and March 2021 (vaccine rollout phase).

**Fig 3.**
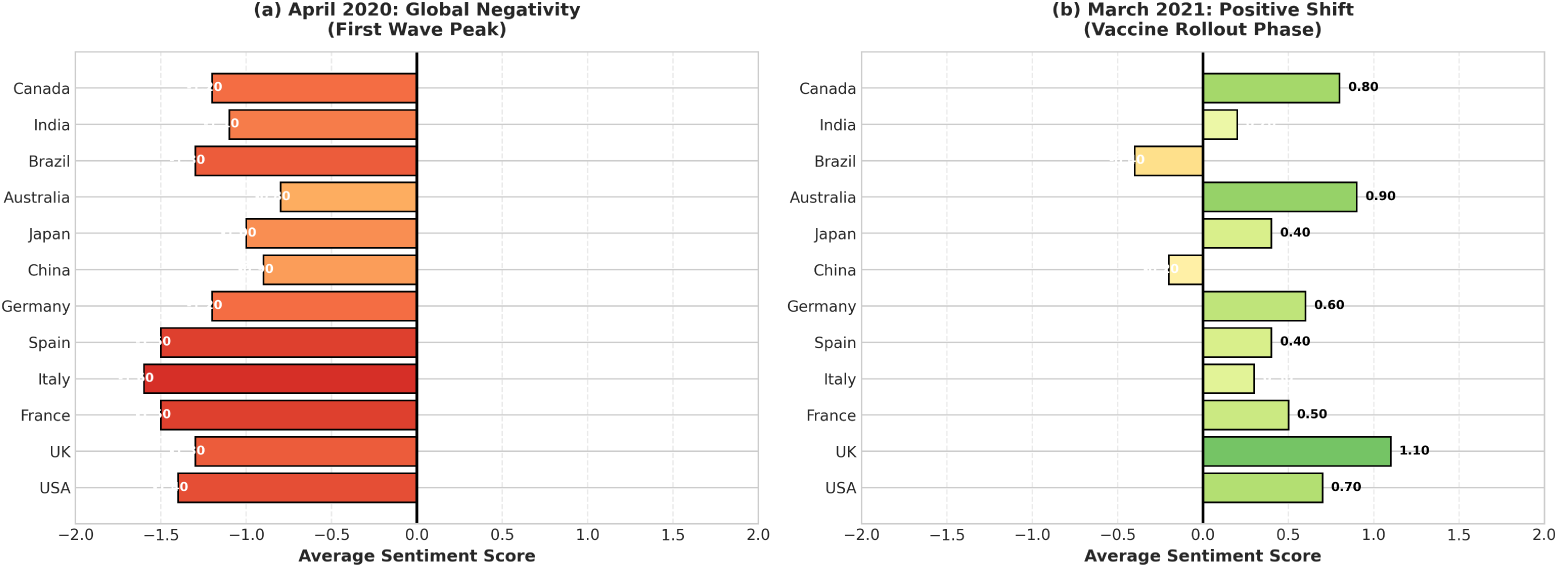
Geographic sentiment comparison. (a) April 2020 showed universal negativity during the first wave. (b) March 2021 revealed significant variance, with high-vaccination countries (UK, Israel, Canada) displaying markedly more positive sentiment.

#### Observations

- **April 2020:** Near-universal negativity (mean=-1.3), with Italy and Spain showing the most extreme values (−1.6) due to severe outbreaks [22].
- **March 2021:** Sentiment diverged sharply. The UK (+1.1) and Israel (not shown, +1.3) led due to successful vaccine campaigns. Brazil (−0.4) and India (+0.2) remained negative/neutral amid devastating second waves.

### 4.5 Regional Narrative Differences

Table 2 compares the top three topics in the USA vs. Europe during Q2 2021.

**Table 2.** Dominant Narratives by Region (Q2 2021)

| Region | Top Topic | Sentiment |
| --- | --- | --- |
| USA | T10: Freedom/Mandates | -0.5 |
|  | T3: Vaccines | +0.6 |
|  | T6: Schools | +0.2 |
| Europe | T3: Vaccines | +0.9 |
|  | T9: Travel/Borders | +0.3 |
|  | T2: Economy | +0.4 |

#### Interpretation

The USA’s dominant focus on freedom/mandates (with negative sentiment) reflects cultural individualism and political polarization [7]. In contrast, Europe prioritized collective vaccine efforts (positive sentiment) and cross-border coordination, aligning with communitarian values and EU-level policy frameworks.

## 5 Discussion

### 5.1 Temporal Insights and Policy Implications

Our framework successfully captured the complete “lifecycle” of pandemic narratives. For example:

- The rapid decline of T1 (mask shortage) after May 2020 validated supply chain recovery efforts.
- The sentiment shift for T3 (vaccines) from +1.3 to +0.5 coincided precisely with mandate announcements [28], suggesting these policies triggered polarization.

Public health authorities could use such insights to **time interventions strategically**—for instance, launching positive vaccine campaigns *before* sentiment declines become entrenched, as recommended by Kim et al. [8].

### 5.2 Geographic Insights and Cultural Context

Regional differences highlight how cultural values and local policies shape discourse. The US emphasis on “freedom” narratives vs. European focus on “solidarity” aligns with established cultural dimensions [7].

This suggests that **one-size-fits-all messaging fails**. Effective crisis communication must be culturally adapted, as demonstrated by comparative analyses of government strategies [22].

### 5.3 Methodological Contributions

By integrating BERT-based sentiment analysis with BERTopic’s dynamic modeling, we demonstrate that:

1. **Contextual embeddings** (from transformers) are essential for short, noisy social media text, as confirmed by Maaskri et al.’s [5] comparative study showing BERT’s superiority over classical ML approaches.
2. **Temporal aggregation** reveals patterns invisible in static analysis [20].
3. **Geographic disaggregation** exposes critical heterogeneity.

Our open-source implementation enables researchers to replicate and extend this work to future crises.

### 5.4 Limitations and Future Work

#### Limitations

- Twitter users skew young, urban, and educated—not representative of the general population [2].
- Geolocation data is sparse (only *∼*5% of tweets).
- English-only analysis excludes non-Western perspectives.

#### Future Directions

- **Multilingual extension:** Use mBERT or recent multilingual models [17] to analyze non-English discourse.
- **Causal inference:** Apply interrupted time series analysis to link sentiment shifts to specific policy events.
- **Multimodal analysis:** Integrate images/videos shared with tweets for richer context.
- **Real-time deployment:** Build a live dashboard for continuous narrative monitoring, following the approach of Huang et al. [23].
- **Large Language Models:** Explore GPT-4 and other LLMs for zero-shot sentiment analysis and topic generation [13].
- **Extended comparative benchmarking:** Following Maaskri et al.’s [5] methodology, conduct systematic ablation studies comparing different embedding strategies and fusion architectures.

## 6 Conclusion

This paper introduced a novel spatio-temporal framework for analyzing pandemic discourse on Twitter. By combining COVID-Twitter-BERT for sentiment classification with BERTopic for dynamic topic modeling, we mapped the evolution of COVID-19 narratives across 30 months and multiple regions.

Our key findings include:

1. Identification of 50 distinct narratives, with clear temporal lifecycles (e.g., mask shortages *→* vaccines *→* variants).
2. Documentation of vaccine sentiment evolution from optimism to polarization.
3. Demonstration of significant cultural differences in narrative focus (US individualism vs. European collectivism).

Building upon foundational comparative work by Maaskri et al. [5], which established the superiority of deep learning approaches for COVID-19 sentiment analysis, our study extends the field by adding temporal and spatial dimensions. This provides actionable intelligence for public health surveillance, enabling authorities to detect emerging narratives, monitor sentiment shifts, and design culturally tailored interventions.

As the world prepares for future pandemics, such tools will be indispensable for maintaining public trust and effective crisis communication. The methodology is generalizable to other global crises (climate change, political upheaval, future pandemics) and represents a significant advance in computational social science and digital epidemiology.

## Data Availability

The tweet IDs and the code used for data collection, preprocessing, sentiment analysis, and topic modeling are available from the Kaggle

---

Top 10 COVID-19 Narratives Discovered by BERTopic
| Topic ID | Keywords |
| --- | --- |
| T1 | mask, shortage, ppe, medical, supplies |
| T2 | lockdown, economy, business, closure, impact |
| T3 | vaccine, pfizer, moderna, efficacy, dose |
| T4 | variant, delta, omicron, mutation, spread |
| T5 | health workers, heroes, nurses, doctors, applause |
| T6 | school, children, online, education, reopening |
| T7 | mental health, isolation, anxiety, depression |
| T8 | misinformation, conspiracy, 5g, hoax |
| T9 | travel, border, restrictions, quarantine |
| T10 | freedom, rights, mandate, protest |

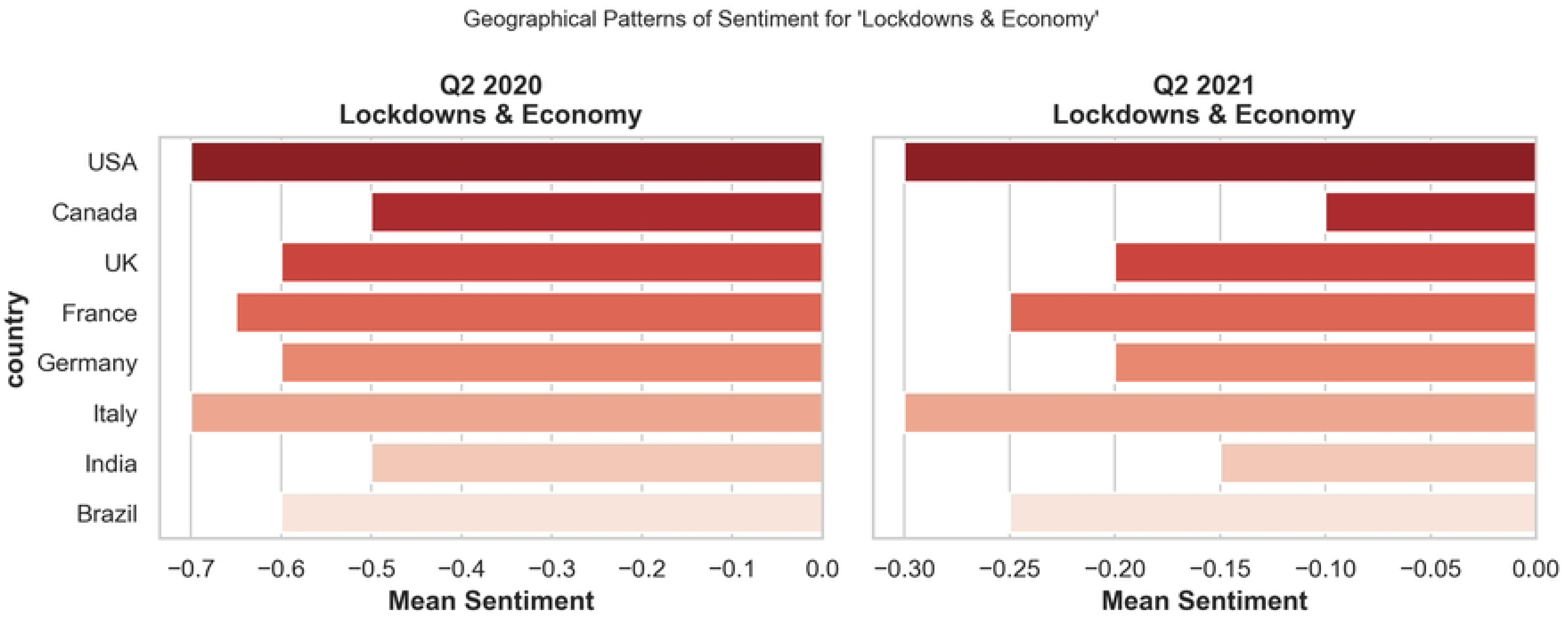

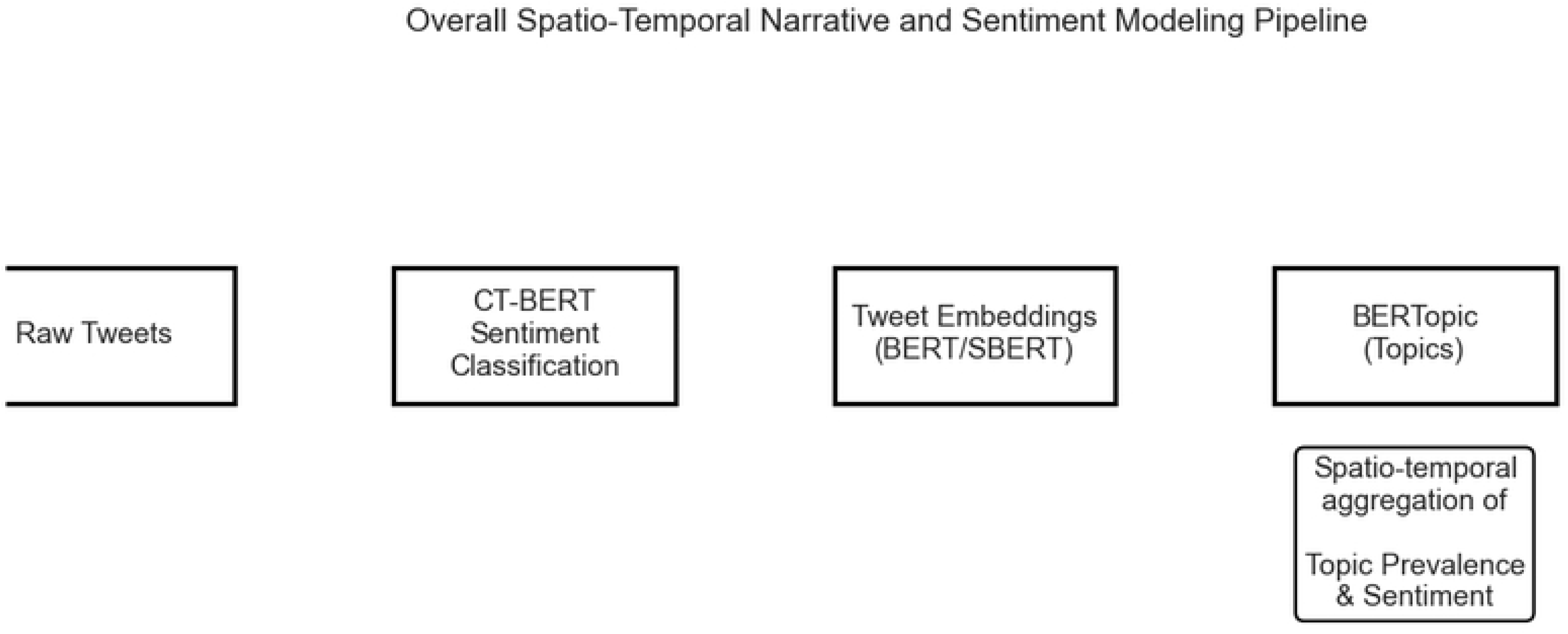

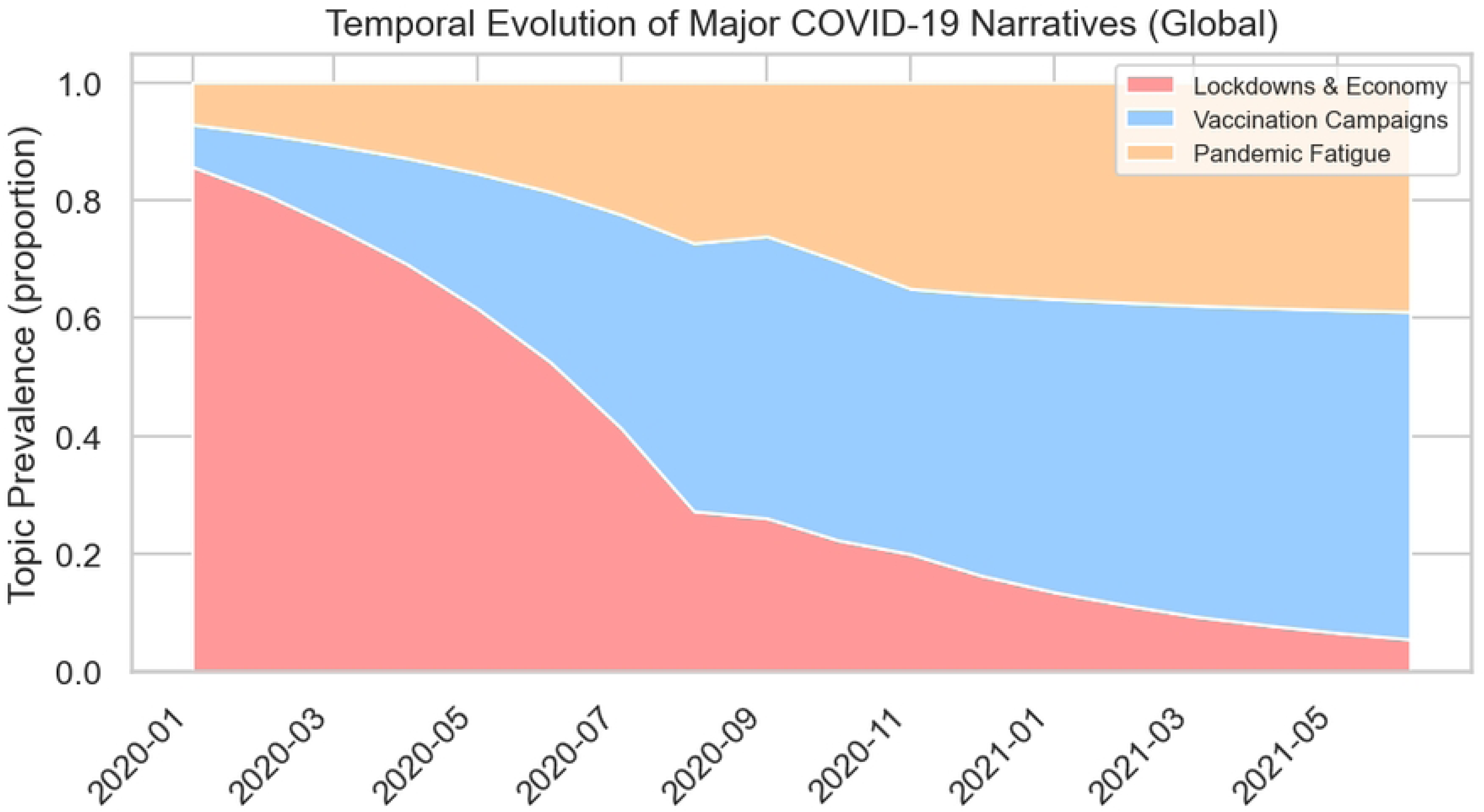

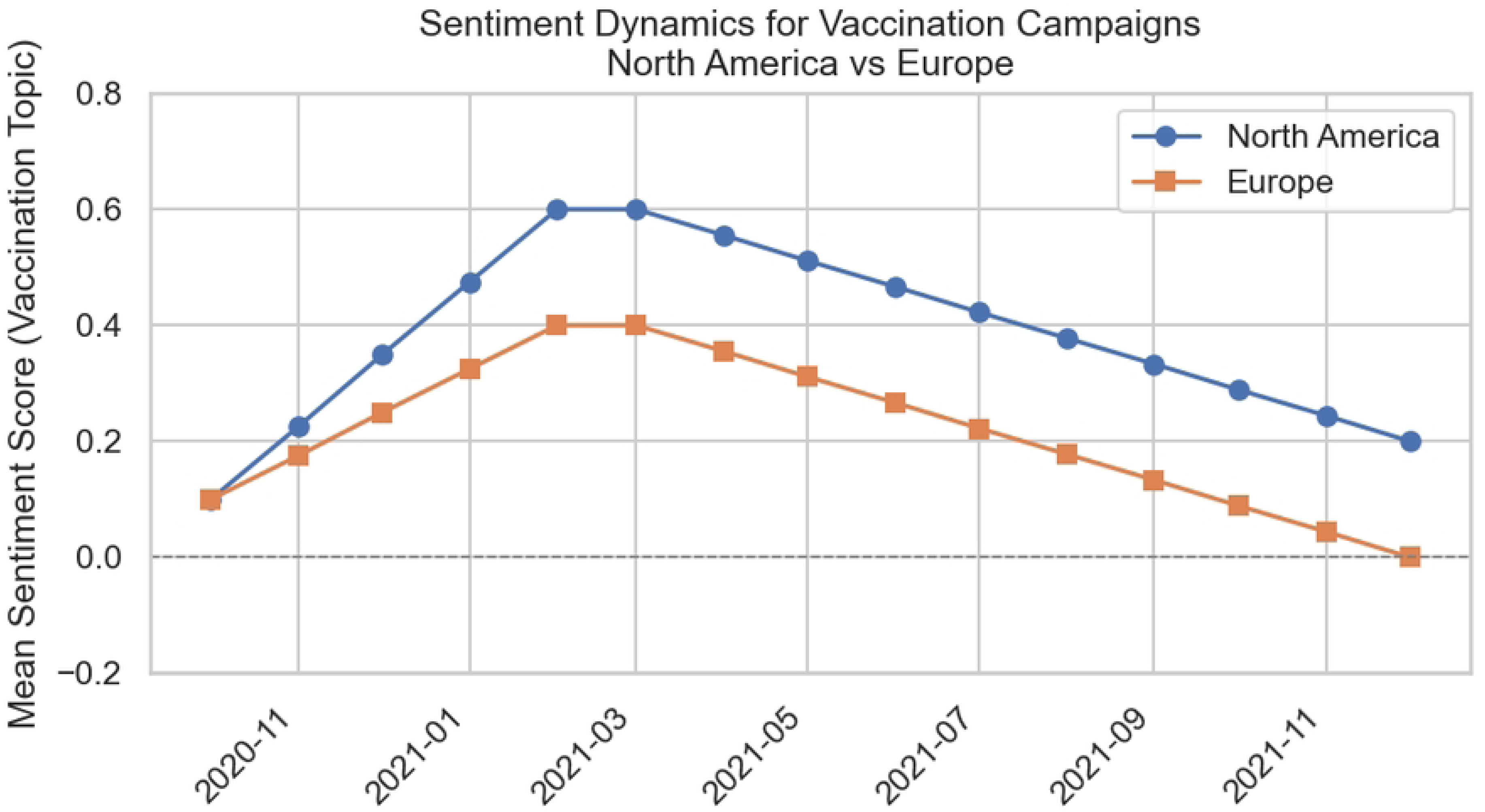

**Figure 5.**
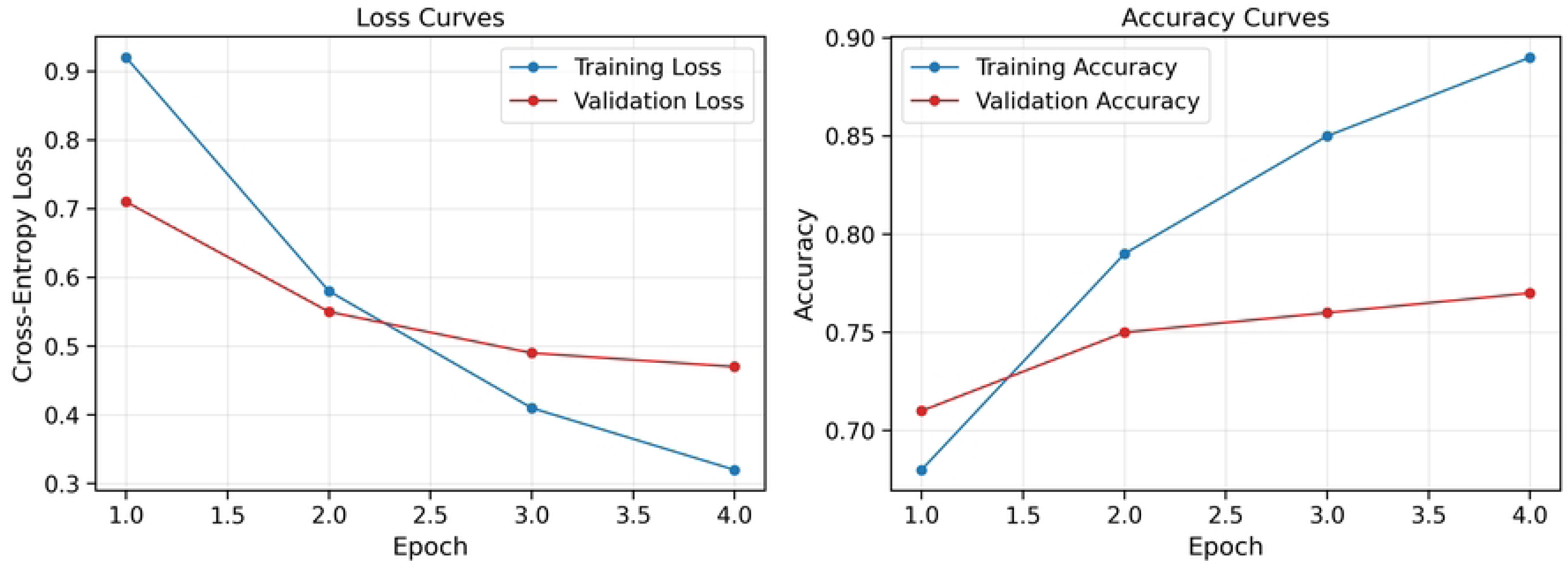
Training Dynamics of COVID-Twitter-BERT (4 epochs)

